# Natural interaction based visual field prediction in virtual reality – a pilot study

**DOI:** 10.64898/2026.10.27.26366299

**Authors:** Samira Frerichs, Melanie Kempf, Björn Severitt, Siegfried Wahl, Alexander Neugebauer

## Abstract

**Objective:** The aim was to develop and evaluate the Passive Visual Field Measurement (PVFM), a novel visual field (VF) measurement tool that predicts the VF from a user’s natural, intuitive interactions in a virtual reality (VR) environment. Due to the flexible nature of its input, the tool could be integrated with various interactive VR tasks and even run passively in the background of tasks users are already motivated to carry out, such as preventive exercises or rehabilitation training.

**Methods:** The PVFM was developed and compared to a conventional VF measurement method, kinetic perimetry using the Octopus 900 perimeter, in a pilot study with n=4 retinitis pigmentosa patients. Both measurements were carried out for each eye, for a total of eight sets. The measured VFs were compared using radial-weighted Intersection over Union (IoU), showing the degree to which PVFM results align with conventional VF testing.

**Results:** For patients with largely concentric visual fields (n=6 eyes), agreement between PVFM and kinetic perimetry was high, with radial-weighted IoU of 75.4% to 91.0% and mean absolute angular distances between VF boundaries of 0.31° to 2.68°. In contrast, one patient with pronounced peripheral visual field areas (n=2 eyes) showed substantially lower agreement, with IoU of 29.6% to 35.7% and distances of 17.5° to 22.3°, as kinetic perimetry detected a larger peripheral visual area than PVFM. PVFM takes significantly longer than kinetic perimetry, however patient feedback indicates that PVFM is more enjoyable and engaging.

**Conclusion:** The preliminary evidence of this pilot study suggests the feasibility of PVFM as an intuitive and engaging method for detecting VF boundaries in patients with concentric peripheral visual field loss, with agreement with kinetic perimetry within the range of test-retest variability reported for conventional kinetic perimetry. However, PVFM showed drastically lower agreement in peripheral, non-concentric visual field areas, a current limitation requiring further investigation and refinement.

## Introduction

Visual field (VF) testing is an essential tool for the diagnosis and continuous monitoring of various retinal and neuro-ophthalmological conditions such as glaucoma, retinitis pigmentosa, and age-related macular degeneration, all of which are characterized by the loss of parts of the visual field, so-called visual field defects. In glaucoma and retinitis pigmentosa in particular, this loss typically starts in the periphery and progresses inward, eventually leaving only a constricted central visual field, and can ultimately end in complete blindness [1, 2]. Crucially, this process is often difficult for affected individuals to notice on their own. Rather than perceiving a clear dark patch or a black tunnel where vision has been lost, as is oftentimes shown in visual representations or simulations of such conditions [3, 4], the visual system tends to perceptually complete these missing regions, and a considerable proportion of patients remain unaware of their visual field loss even when directly asked about it [5]. As a consequence, these conditions are often not diagnosed until severe, irreversible damage has occurred, and even after diagnosis, regular visual field testing remains essential to detect and respond to further progression.

A conventional method for VF testing, especially in patients with extensive peripheral vision loss, is kinetic perimetry [6–9]. Patients are tasked with fixating a central marker on a screen in front of them while a target moves from a non-seeing area of the visual field towards a seeing area (e.g., from the non-seeing periphery towards the seeing center). As soon as the target is perceived, the patient presses a button, and the target’s position at that moment – oftentimes corrected by reaction time - marks a point on the boundary of the visual field. By repeating this process from different directions and with varying start and end points, the boundary of the patient’s visual field can be reconstructed by connecting the individual points. The result is known as as an isopter field, where an isopter line describes the connecting line between two adjacent boundary points [7].

While kinetic perimetry continues to be the convention for clinical visual field testing, it comes with notable limitations. It is typically performed by a trained medical professional using stationary, high-cost perimeters, which makes testing only feasible within clinical settings and makes it costly and logistically demanding to repeat frequently [10, 11]. Since routine clinical visits are oftentimes scheduled multiple months or even years apart, sudden changes in a patient’s condition can go undetected for extended periods, which reduces the likelihood of identifying disease progression early enough to intervene effectively.

These constraints have led to a growing interest in home-based, accessible, and frequent VF monitoring, particularly for patients with chronic and progressive conditions such as retinitis pigmentosa and glaucoma [12, 13]. Advances in virtual reality (VR) technology have made this increasingly feasible. Modern head-mounted displays are self-contained and portable and offer a considerably larger field of view than conventional monitor-based setups with common visual field sizes of 90-110° diameter [14]. They also provide head-tracking, which either enables the user to freely look around a virtual scene or, when head-centric fixation is required, to display visual stimuli at head-fixed positions, unaffected by accidental head movements. Building on these properties, a growing number of VR-based visual field testing devices have been developed in both academic and commercial contexts [10], such as the Advance Vision Analyzer (AVA) (Elisar VisionTechnology) [15, 16], VisuALL (Olleyes) [17], and the Toronto Portable Perimeter [18], several of which show good agreement with the current clinical standard for visual field testing [18–21]. However, a two-year home-monitoring study, using the Toronto Portable Perimeter, found that patient compliance declined significantly over the course of the study, indicating that simply making a device available at home does not by itself guarantee sustained, motivated use [18]. These previous findings suggest that frequent VF screening at home is feasible, while also highlighting the importance of user motivation and engagement.

This points to a second limitation of conventional perimetry: it can be mentally and visually demanding. Continuously fixating a single marker for several minutes – while also staying alert to press the button once the peripheral stimulus becomes visible - is cognitively taxing and can lead to fatigue [22], which in turn can lower patient adherence and reduce the reliability of the resulting measurements. This becomes especially problematic for patients who find it difficult to sustain attention on a single task for an extended period, such as young children or individuals with cognitive impairments [23, 24].

One method developed to address this issue is Saccadic Vector Optokinetic Perimetry (SVOP) [25]. Rather than requiring patients to maintain fixation and press a button, SVOP uses eye tracking to detect natural, involuntary saccadic eye movements towards stimuli, removing the need for sustained fixation or a manual response and making testing considerably more comfortable for children and other patients who struggle with conventional perimetry [25]. More recent VR-based methods use a similar approach, integrating eye-movement-based VF testing in head-mounted displays and reporting that such VR-based versions were consistently preferred by patients over both conventional perimetry and screen-based eye-tracking equivalents [26]. A limitation of these approaches, however, is that they depend on VR headsets with integrated eye-tracking hardware, which remains considerably more expensive than standard consumer VR headsets and is, for the most part, still confined to devices aimed at research, enterprise, or industry use rather than the casual consumer market.

Beyond diagnostic assessment, VR technology has also shown promise in other areas of visual healthcare, notably visual rehabilitation. Several home-based, VR-delivered training programs have been developed for patients with visual field defects, with reported improvements in visual perception and quality of life [27, 28]. Since patients in such rehabilitation tasks already interact naturally with stimuli distributed across the device’s field of view, this raises an opportunity: the way a patient searches for and responds to these stimuli may itself carry information about the size and shape of their visual field, without the need to administer a separate, dedicated VF test.

This work is motivated by this possibility. We propose a novel Passive Visual Field Measurement (PVFM) method for VR that estimates a user’s visual field purely from the position and timing of their natural search-and-click behavior towards targets distributed throughout the virtual scene, without requiring the patient to fixate a specific marker and without a need for eye-tracking integration. Because the method has no fixed requirements regarding the visual scene beyond the presence of variably positioned targets to interact with and an incentive to find them quickly, it can, in principle, run passively in the background of a variety of gamified VR tasks, including visual rehabilitation exercises. This would allow to combine visual field monitoring and visual rehabilitation within a single, interactive and accessible VR experience, enabling patients to track changes in their visual field over time as an unobtrusive byproduct of tasks they are already motivated to perform.

In a pilot study, we compare the PVFM method against a kinetic perimetry VF test in a group of patients with peripheral visual field defect caused by retinitis pigmentosa. The comparison is not intended as a test of medical validity or to replace standard kinetic perimetry for clinical diagnostics, but rather as a feasibility and design study with the goal of exploring whether VR based, eye-tracking-free visual field testing can provide patients with a simple and accessible way to monitor their own visual field.

## Methods

This research work focuses on two objectives:

1. The design and development of the PVFM tool.
2. The experimental comparison between the PVFM tool and conventional kinetic perimetry in a patient study.

### Development of the passive visual field measurement

The PVFM tool was developed using the Unity 3D engine (Version 6000.2.6f2) and was integrated with a VR based search task developed in the same engine. The visual task and the PVFM tool are closely connected, but each focuses on a different aspect: The visual task is designed to encourage interactions with the virtual scene, whereas the PVFM tool uses the input data of these interactions – time and relative positions – to determine visual field boundaries.

#### Visual task design

The visual task features a single target, positioned randomly within a specified spawn area in front of the patient (Fig 1). Users are instructed to search the area for the target and – once they spot it – select it through point-and-click action with the VR controller. As soon as the target is selected, it disappears and a new target immediately spawns at a different position, repeating the process.

**Fig 1.**
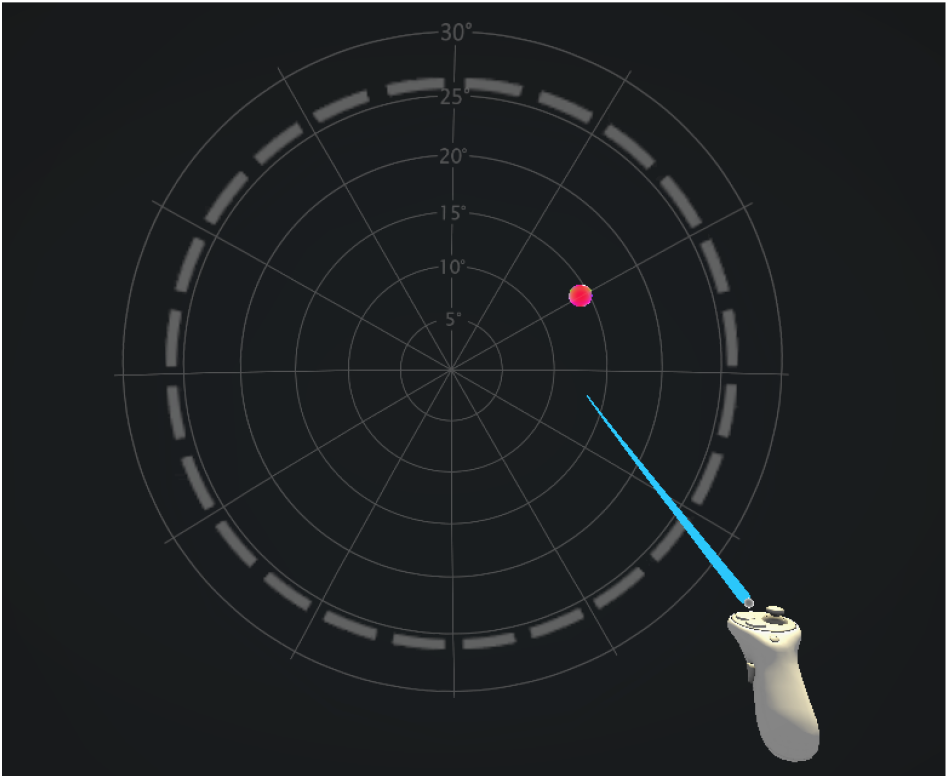
Visual task layout. Visualization of the circular spawn area (dotted white line), search target (red dot) and controller with selection ray. The radial scale (fine white lines) was not visible during the experiment and is only used to visualize the size of the spawn area.

The spawn area – the area in which the targets may appear - is a circular, spherically curved field at four meters distance from the user’s viewpoint in the virtual environment. Its size *R*_spawn_, i.e. the maximum eccentricity from the patient’s forward-facing direction at which targets could appear, could be set manually in advance of the session. During the experiments, this feature was used to prevent patients with small visual fields from having to search a large area, which would increase the average search time drastically.

Each session comprised a minimum of 360 samples, where one sample describes one target being selected. The position in which new targets would spawn was limited not only to the spawn area, but also to a maximum distance to the previous target. This maximum distance was set to be equal to *R*_spawn_. This limitation was implemented as samples with higher distance between two targets would be less relevant for visual field measurement, as will be explained in the following chapter.

Each eye can be tested individually by displaying the visual content within the VR headset only to the left or right screen, respectively. Additional optimizations to the target spawn behavior, utilizing runtime feedback from the PVFM tool, were integrated, further explained in the Chapter Visual task optimization techniques. First, however, we will focus on the design of the PVFM tool itself.

#### Passive Visual Field Measurement Tool

The aim for the PVFM tool is to predict the visual field of a patient based on only their interactions within the VR scene, without forcing patients into unnatural or non-intuitive behavior such as having to fixate the gaze on a central marker. Thus, the calculation of the visual field must be based on parameters that can be extracted from such natural interaction alone. Specifically, for the PVFM, two interaction parameters are used: Relative position between two consecutive interactions, and time between those two interactions. The relative position is further divided into two separate parameters – the direction and the magnitude of the vector between the two targets. Given the spherical nature of visual field measurements, where distances are usually measured in visual degree rather than any linear unit of length, we will refer to the two positional parameters as radial angle for the direction and angular distance for the magnitude of the visual angle between the two. Lastly, the time between two interactions will be referred to as response time. Figure 2(a) shows a visual explanation of the radial angle and angular distance parameters.

**Fig 2.**
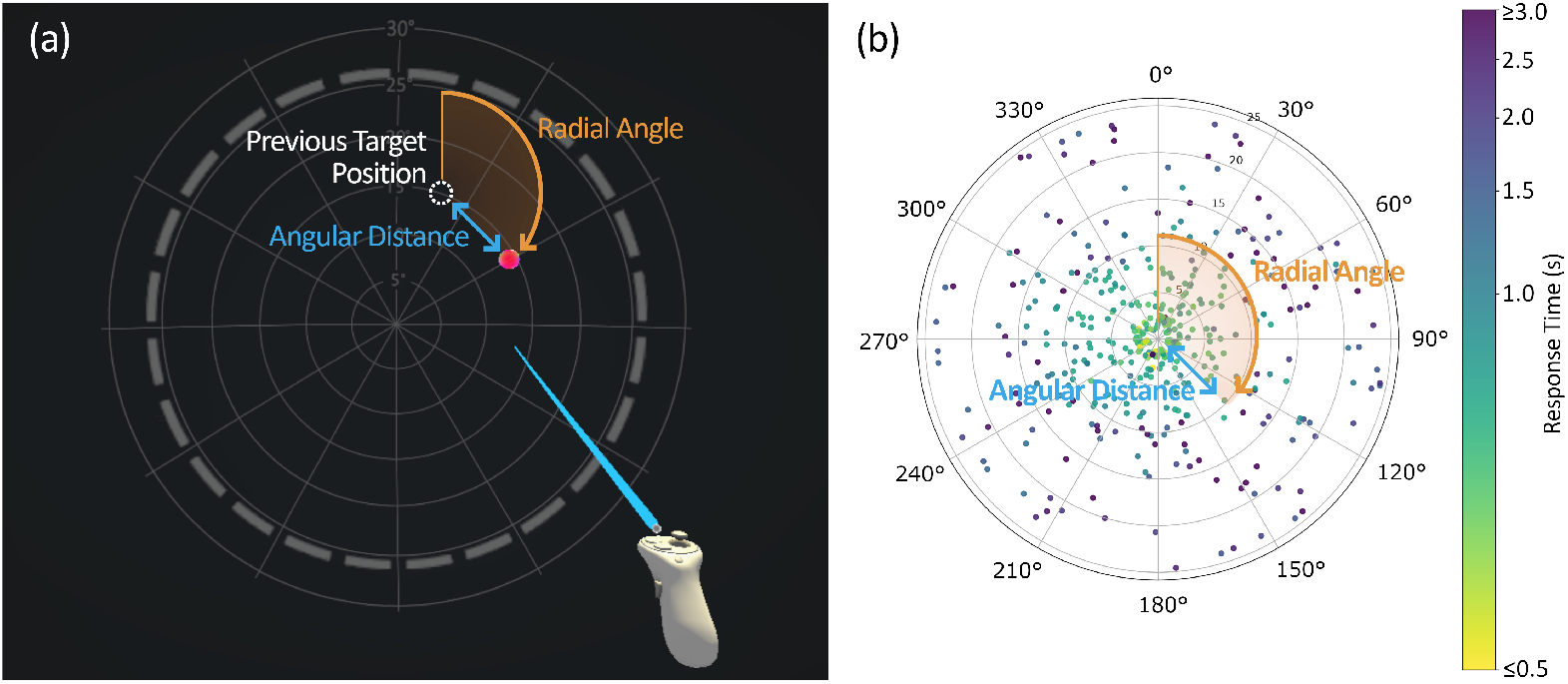
Sample parameter visualization. (a) Visual representation of the two positional parameters within the visual search task: The radial angle, describing the direction of the vector between previous and current selected target, and the angular distance, describing the magnitude of the visual angle between them; (b) Representation of the same sample normed to the center of a radial graph, together with all other samples of one measurement.

With each interaction in the visual task, a sample is recorded, containing the radial angle, angular distance, and response time between this and the previous interaction. Figure 2(b) shows an example of this and other samples of the measurement being normed to the same center point and colored based on response time, resulting in a map of response times for different eccentricities in the user’s visual field. This representation may already give an idea of how a visual field could potentially be predicted. The core concept of the PVFM algorithm to predict the VF builds on the following assumption: If, after an interaction with one target, the next target is already within the patient’s VF, the response time will likely be short, since the patient can directly focus and select the second target without having to search for it. If the next target is at a position that is not within the patient’s VF, then the patient first has to scan for the second target, significantly increasing the expected response time. Providing that, the VF area will be distinguished by samples with significantly shorter response times than the samples outside the VF. Thus, the response time of samples at different positions in the tested field of view may provide sufficient information to predict the VF.

As stated in the first Chapter Introduction, the results of a kinetic perimetry measurement are commonly represented in the form of an isopter field, an approximate outline of the VF area [7]. This representation makes the VF easy to visually interpret and is therefore one of the standard representations in clinical visual field diagnostics. Thus, the aim for the PVFM is to provide such an isopter field. This poses a challenge, because unlike kinetic perimetry, PVFM does not provide inherent boundary points of the visual field. However, given the previous assumption regarding response times being significantly shorter within the VF than without, it can be assumed that the boundaries of the visual field mark a steep transition from short to long response times. Thus, by finding where these transitions happen, the VF boundaries can be predicted. In the following, we will explore the method applied.

In a first step, all samples of one measurement set (representing all samples for one eye in one patient) are divided into sections – a process called “binning” - based on their radial angle. Specifically, each sample is assigned to the closest 30° meridian, e.g. samples from -15° to 15° are assigned to the 0° meridian, samples from 15° to 45° are assigned to the 30° meridian, and so forth. The choice of binning samples into twelve 30° segments was made as it is in line with the 8-16 meridians commonly tested in standard kinetic perimetry [8, 29]. A higher number of segments would increase the number of required samples whereas a smaller number would reduce the accuracy of the visual representation of results.

Next, samples in each bin are sorted based on their angular distance. By going through all samples from smallest to largest angular distance and assessing the response times for each one, it can be assumed that there will be a noticeable change in response time at the angular distance where the seeing field transitions into a non-seeing field – i.e. where the boundary of the visual field is. Similarly, peripheral windows of remaining vision will be characterized by a decrease in response time at the eccentricity of the inner (more central) edge of the window, followed by yet another increase at the outer edge. The algorithm to determine these boundaries for a given meridian is visualized in Figure 3.

**Fig 3.**
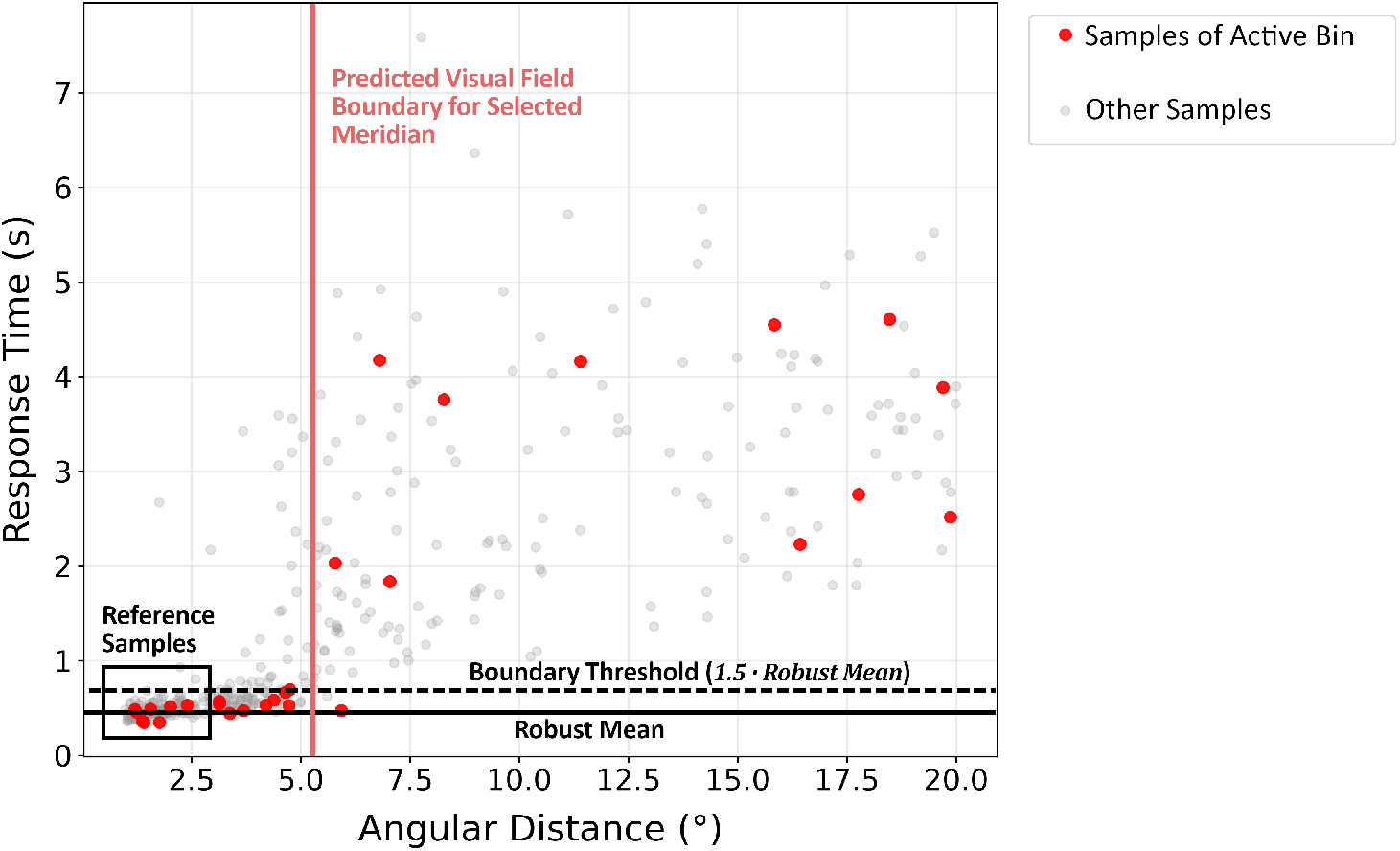
Visual field boundary detection. Visualization of the algorithm to determine the boundary of a patient’s visual field. As each meridian of the visual field is analyzed individually, samples of one such meridian are marked in red.

The algorithm uses threshold-based boundary detection. For each of the 12 meridian segments, it selects the five samples closest to the center (i.e., with the smallest angular distance), resulting in 60 reference samples in total. As will be explained in the following chapter , these samples can be considered to be within the user’s visual field. From these reference samples, a robust estimate of response time is computed using the median and the median absolute deviation (MAD). The MAD is defined as the median of the absolute deviations between each sample’s response time and the median response time of the set [30].

A cutoff threshold *t*_cutoff_ is then defined as:

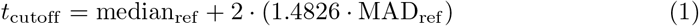

The factor 1.4826 rescales the MAD so that it is comparable to the standard deviation under a normal distribution [31], while the factor 2 was chosen following the literature convention for a slightly sensitive cutoff multiplier [32].

Any reference sample with a response time above this threshold is capped at the threshold value, limiting the influence of individual outliers. The mean of the now capped reference samples is then calculated, yielding a robust mean response time.

This median/MAD-based approach was chosen over a simple mean-and-standard-deviation threshold as it is substantially more resistant to outliers. Since this robust mean is calculated from reference samples that are certain to be within the VF, it represents the baseline response time assumed for samples within the VF. From this baseline, a boundary detection threshold *t*_boundary_ is calculated as:

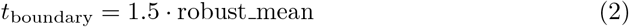

The decision to make the boundary detection threshold a simple factor of the robust-mean, rather than using a more conventional variance-based threshold, will be elaborated in the discussion in Chapter Learnings from the acquired data. Consecutive samples that fall consistently above the threshold are considered to be outside the VF, whereas consecutive samples under the threshold indicate VF areas. Specifically, samples of each meridian are analyzed in order of eccentricity, starting at the most central sample. If a sample’s response time lies above the threshold, it is considered a potential candidate for a visual field boundary. To confirm, a second condition must be met: The median of the next five samples in order must also be above the threshold, ensuring that the current sample is not just an outlier. If both conditions are met, a boundary line from seeing to non-seeing area is marked at the average angular distance between the last sample considered within the VF and the first sample considered without the VF (see ‘Predicted Visual Field Boundary’ in Figure 3). The window size of five samples were selected empirically during initial testing, as a compromise between robustness to noise and sample size limitations. The median window must be large enough that isolated outliers do not trigger a false boundary. A larger window, however, would become impractical given the limited number of samples per meridian. An odd value was chosen so that the median is always a single, unambiguous sample value.

It is possible - in case of non-concentric visual fields or windows of visual field rest – that there are multiple visual field boundaries along a meridian. Thus, the analysis of samples’ response times along their order of eccentricity is continued even after finding an initial boundary. When at least three consecutive samples fall below the threshold, the average eccentricity between the last sample above the threshold and the first of the samples below the threshold is marked as a boundary point between non-seeing and seeing area, and the initial conditions for finding a boundary from seeing to non-seeing area apply once again. Again, this condition of requiring three consecutive below-threshold samples was selected empirically during initial testing. It needs to avoid false boundary detection from accidental fast responses (which can happen quite regularly if the patient happens to move their gaze in the right direction by chance) while still allowing narrow islands of preserved vision to be detected.

Once all predicted boundary points have been calculated for each of the twelve meridians, the isopter field – representing the predicted visual field of the patient – can be created by connecting boundary points of adjacent meridians. This provides the desired visual representation of the visual field analogous to the results of kinetic perimetry.

An additional adjustment factor was added, increasing the threshold with increasing eccentricity. This addition was motivated by two known effects on response time: First, reaction time naturally increases with the eccentricity of the stimulus - approximately at a rate of 1.8*ms/deg*_eccentricity_, as determined in preliminary studies such as by Schiefer et. al. [33]. Second, response time is also affected by the time needed to move and orient the VR controller, which likewise increases with angular distance. The combined effect of eccentricity on response time was approximated for each individual patient. The factor by which the response time threshold increases is calculated as simple linear regression between the angular distance as independent variable and response time as dependent variable, based on the first 10 samples of each meridian in both eyes. Outlier samples not within the MAD-based capping-threshold described in Equation 1 were not included for regression calculations. This eccentricity-based threshold adjustment was applied to increase the accuracy of the detected visual field boundaries, in particular for peripheral visual field rests where the effect of an eccentricity-adjusted threshold is strongest. This factor was calculated only for the final boundary detection, as it relies on sufficient reference samples for the estimate.

#### Visual task optimization techniques

In order to improve the information attained from a limited number of samples, some optimizations have been introduced to the task design.

By default, new targets are spawned at random angular distance *d* to the selected target, with the maximum angular distance equal to the spawn area radius *R*_spawn_:

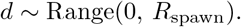

However, to ensure a sufficient number of reference samples for each meridian, half of the first 10 samples in each 30° segment are guaranteed to spawn at a very low angular distance to the previous sample — lower than half their predicted VF size —, ensuring that the sample is within the patient’s VF when spawning.

In addition, to increase the accuracy of the measurement with limited sample size, a method to optimize the spawn positions of samples was implemented: Once at least 15 samples were recorded for a specific meridian, the visual field boundary prediction described above was carried out with each following new interaction. If one or multiple boundaries were found for this meridian, half of all following samples within this segment are adaptively spawned in a gaussian distribution around a predicted boundary distance *R*_boundary_:

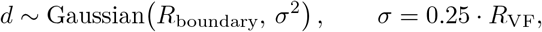

where *R*_VF_ is the predicted visual field radius. This technique aims at increasing the sample density in areas of high interest, i.e. near the visual field boundaries.

Lastly, the number of samples per measurement could extend beyond the defined minimum of *N*_min_ = 360 when not all meridians had at least one predicted boundary larger than 0^°^. This condition was introduced since boundary predictions would occasionally fail in some meridians due to a low number of samples in the periphery.

### Experimental evaluation

To investigate the feasibility of the PVFM method as a monitoring tool, specifically for peripheral visual field defects, a patient study was carried out. The study aimed to compare the VF predicted by the PVFM to the VF measured using conventional kinetic perimetry as well as to gather patient feedback on the usability of and experience with the PVFM tool.

#### Ethics

The study was approved by the ethics committee of the faculty of medicine and the university hospital of the University of Tübingen (Nr. 701/2024BO1) in accordance with the 2013 Helsinki declaration. Patients were recruited and measurements were taken in the time from May 1 to May 31, 2026. All participants received and signed written informed consent forms.

#### Study population

Four patients, diagnosed with retinitis pigmentosa and a resulting peripheral visual field defect, were recruited for this study. Patient details as well as prior experience with navigation aids and VR devices, acquired through an initial questionnaire at the start of the session, are summarized in Table 1. Visual Acuity of both eyes was measured by the experimentor at the start of the session using the ZEISS VISUSCREEN digital Snellen chart system (ZEISS) [34].

**Table 1.** Patient data.

| Patient | Age, Sex | Years since diagnosis | Visual acuity (dec.) (LE / RE) | Navigation aids | VR experience | Vision correction |
| --- | --- | --- | --- | --- | --- | --- |
| 1 | 66-70, f | 31-35 | 0.80 / 0.40 | - | Some | Glasses |
| 2 | 46-50, f | 11-15 | 0.32 / 0.20 | White cane | - | - |
| 3 | 61-65, f | 41-45 | 0.32 / 0.32 | White cane | Some | Glasses |
| 4 | 21-25, f | 6-10 | 0.40 / 0.50 | - | Some | Glasses |

Patient demographics and clinical background.

Patients were recruited through a mailing list. Two patients had previously taken part in a different study on visual field defects at the facility and had agreed to be contacted about future research opportunities. The remaining two patients had subscribed to the mailing list independently to stay informed about opportunities to participate in VR studies.

### Study design and procedure

The study consisted of one session of 120 minutes per patient. After the introduction and filling of the initial questionnaire, each patient completed one kinetic perimetry test and one PVFM test for each eye. For patients 1–3, kinetic perimetry was performed after PVFM; for patient 4, this order had to be reversed due to limited availability of time slots with the kinetic perimetry device. After the measurements, a second questionnaire about the patient’s perception of both methods was filled.

#### Kinetic perimetry procedure

Kinetic perimetry was performed with an Octopus 900 perimeter (Haag-Streit), using the semi-automatic kinetic perimetry setting with a stimulus with the III4e standard (0.43° diameter, 318 cd/m^2^ luminance), moving at a constant angular velocity of 5°/second and in front of a 10 cd/m^2^ luminance grey background. These settings are consistent with those commonly used on this device in clinical context [35, 36]. Both eyes were measured individually. The perimetry was carried out by the experimenter under supervision of a medical professional.

#### Passive Visual Field Measurement procedure

PVFM was performed using a Meta Quest Pro VR headset (Meta) [37]. After a brief introduction to wearing the headset and interacting with the VR controller, patients started the measurement process themselves by pressing a start button in the virtual environment. Again, both eyes were tested individually.

The spawn area’s size *R*_spawn_ was set individually for each patient prior to measurement, based on the visual field size reported by the patients themselves or by previous medical diagnostic reports provided by the patients. Both parameters were set to approximately three times the reported radius, since larger spawn areas would require extensive search in patients with small remaining visual fields, substantially increasing measurement time with no benefits to the results. For patient 1, who reported residual peripheral islands of vision that made a single representative eccentricity difficult to estimate, *R*_spawn_ was set to the maximum supported by the headset’s field of view at 50° radiu1s [14]. For patients 2, 3, and 4, *R*_spawn_ was set to 12°, 25°, and 20°, respectively.

The virtual environment’s background was set to a uniform dark grey (RGB values of (26, 26, 26) at linear scale), corresponding to 10% of the Meta Quest Pro’s stated maximum display brightness of 100 cd/m^2^ [37], and thus approximating a background luminance of 10 cd/m^2^ to match the background luminance of the Octopus 900 perimeter. As will be elaborated in the discussion of this work, the targets could not be set to fully match the appearance of the kinetic perimetry due to missing display brightness. Instead, they were rendered as bright red, bubble-like spheres of 2° diameter, chosen for high visibility and a gamified appearance.

#### Questionnaires

Two questionnaires were administered per patient. The first, completed before testing, recorded demographic information, clinical background, and prior experience with navigation aids and VR devices (Table 1). The second, completed after both VF tests, consisted of four parts: A 10-point Likert-scale assessment of the subjective experience of kinetic perimetry, an equivalent Likert-scale assessment of PVFM, and a set of forced-choice questions in which patients indicated their preferred method across different categories. The full questionnaires and corresponding patient responses are reported in Chapter Questionnaires.

## Evaluation

Data from the experimental study was evaluated in Python (version 3.13.14) using the NumPy (version 2.5.2) and Matplotlib (version 3.11.1) libraries.

A total of eight kinetic perimetry results and eight PVFM sample sets were acquired during the experiment, one of each per patient per eye. The Octopus 900 perimeter used for kinetic perimetry provides a visual representation of the visual field, in the form of an isopter field as described earlier, as part of its standardized measurement procedure. As such, no additional analysis methods were required for these results. For the PVFM data sets, visual field boundaries were predicted using the evaluation process described in the previous Chapter Passive Visual Field Measurement Tool. As mentioned there, eccentricity-based adjustments were applied to the boundary detection threshold during post-experimental evaluation, compensating for the natural increase in response time toward more peripheral interaction targets.

These evaluations provide a visual representation of the visual fields (VFs) measured with both methods, allowing for a direct visual comparison. For a more quantitative comparison, however, the Intersection over Union (IoU) was calculated. IoU describes the ratio between the intersection and the union of two areas and is a common metric for evaluating the similarity between two shapes. Two different IoU variants were applied to the results.

A pixel-wise IoU compares the image output of the two VF areas pixel by pixel, dividing the number of pixels *n*_intersect_ shared by both VF areas by the number of pixels *n*_union_ contained in at least one of them, so 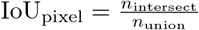. While this represents the most common, cartesian-coordinate based IoU approach, it can be argued that it does not adequately reflect the nature of VF data - since VFs are inherently radial in structure, pixel-wise IoU disproportionately weights peripheral regions, which cover a much larger pixel area than central regions of equal angular extent. For this reason, VF areas are typically reported as square degrees [38, 39], using a radial rather than cartesian coordinate system.

To accommodate this, a second, radial-weighted (or square-degree-wise) IoU was calculated. Here, pixels are weighted by a factor 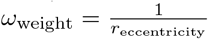 , ensuring that each square degree (defined as 1°_radialAngle_ 1°_angularDistance_) has the same influence on the IoU. An alternative way to conceptualize this radial-weighted IoU is as a measure of the average difference in radius length between the two VFs along any given meridian.

Both IoU methods have merit for comparing the two VF results, depending on the context in which the results are interpreted. Pixel-wise IoU allows comparison of the raw area in which any vision exists, independent of its position within the field of view, while radial-weighted IoU more accurately reflects the comparison of actual perception, as well as changes or differences in VF size.

In addition to the two IoU methods, the mean of all absolute angular distances between the isopter boundaries of the two methods, *d*_meanDistance_, was calculated. This metric quantifies how closely the two boundary lines match in absolute terms, expressed in degrees of visual angle. It is defined as:

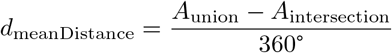

where *A*_union_ and *A*_intersection_ denote the union and intersection areas of the two VFs, respectively, measured in square degrees. The numerator corresponds to the area where the two boundaries disagree, which, when divided by the full 360° radial angle range, results in the average angular distance between the isopter lines of both fields across all meridians.

The low number of samples in the questionnaires (n=4) made a statistical analysis of their results unfeasible. Results are instead presented as raw data.

## Results

### Comparison of visual field measurements

Table 2 summarizes the measurement process for both kinetic perimetry and PVFM, including the number of samples acquired and the duration of each measurement.

**Table 2.** Measurement session data.

| Patient | Number of samples in PVFM | PVFM spawn area radius | PVFM measurement duration (LE/RE) (in min:s) | Kinetic perimetry measurement duration (LE/RE) (min:s) |
| --- | --- | --- | --- | --- |
| 1 | 390/398 | 50° | 14:48 / 17:58 | 11:08 / 5:39 |
| 2 | 396/376 | 12° | 18:13 / 21:45 | 8:45 / 3:54 |
| 3 | 383/396 | 25° | 9:07 / 8:26 | 6:32 / 1:54 |
| 4 | 396/396 | 20° | 10:17 / 9:39 | 1:44 / 1:36 |

Information on the measurement process of kinetic perimetry and Passive Visual Field Measurement (PVFM). Number of PVFM Samples describes the number of target interaction samples acquired per patient per eye. PVFM spawn area radius describes the radius $R_{\text{spawn}}$ of the area in which targets in the visual search task would appear, and simultaneously the maximum angular distance between two consecutive targets. The Measurement Duration values of the two methods show how long the VF measurement for each individual eye took.

It can be noted that the duration of the kinetic perimetry VF measurement was noticeably shorter than the duration of the PVFM (average 5:09±3:18 min and 13:47±4:46 min, respectively). While durations of the kinetic perimetry can vary substantially based on the complexity of the shape of a patient’s visual field as well as the speed of the medical professional carrying out the measurement, the durations in this experiment align with commonly stated durations for kinetic perimetry of 5-10 minutes [40].

Figure 4 shows the visual fields measured with both methods for all patients and eyes, alongside the corresponding IoU values. Results show that the overall agreement between PVFM and kinetic perimetry was strongest for patients with largely concentric visual fields. For patients 2–4, radial-weighted IoU ranged from 75.4% to 91.0%, indicating good alignment between the two methods in these cases. This trend also shows in the low mean absolute angular distances between the isopter lines of both fields, ranging from 0.31° to 2.68°. In contrast, patient 1 showed large peripheral visual field areas in the kinetic perimetry measurement that were largely absent from the corresponding PVFM results. This results in a substantially lower radial-weighted IoU of approximately 30-35% for this patient, with mean absolute angular distance of 17.5° to 22.3°. Potential reasons for this discrepancy will be addressed in the discussion, Chapter Comparison of visual field measurements.

**Fig 4.**
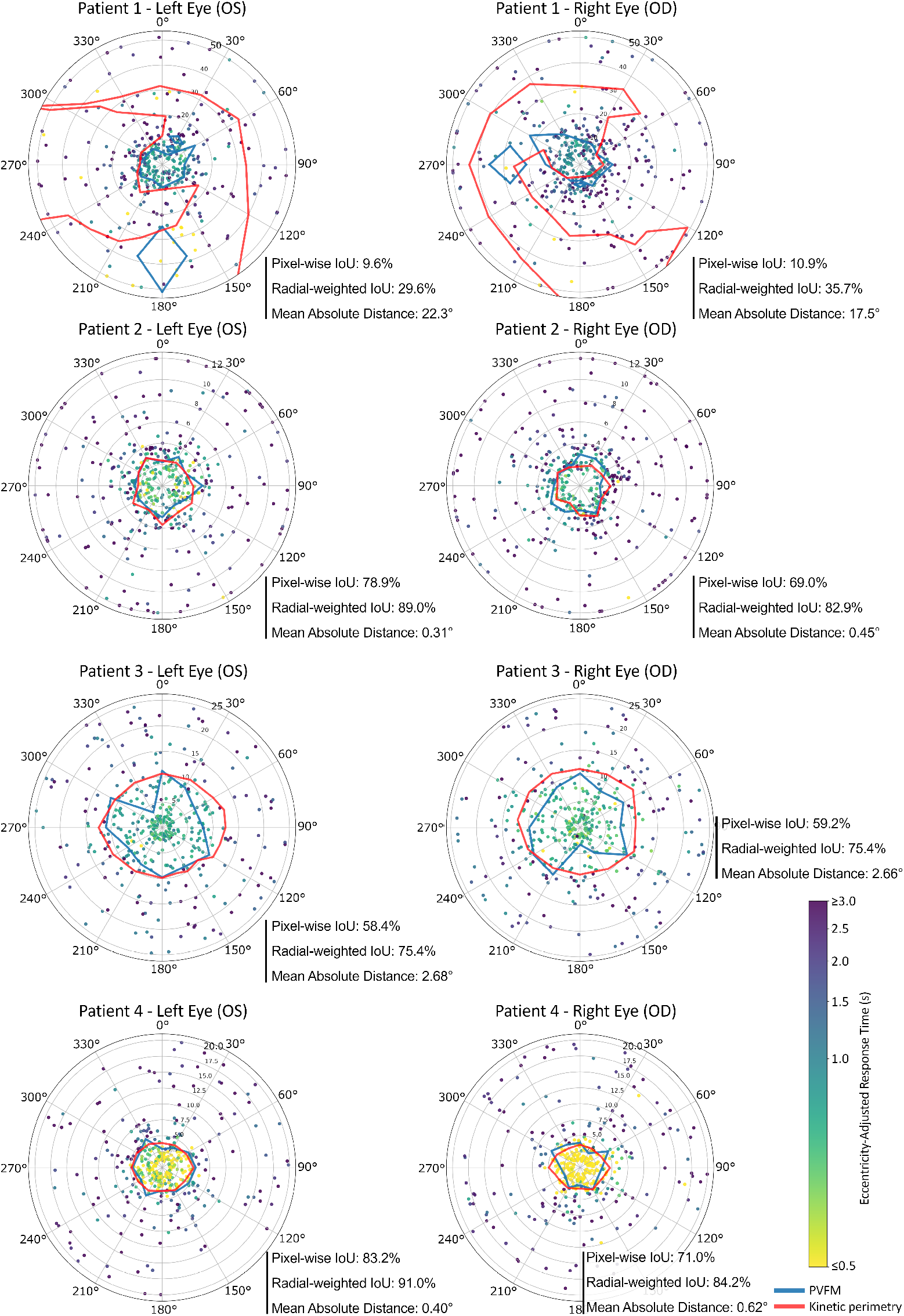
Visual field comparison. Visual field visualization of the PVFM and kinetic perimetry for both eyes. Pixel-wise IoU and radial-weighted IoU quantify the areal overlap between the two visual fields, while the mean absolute angular distance indicates the average radial offset between the two VF boundaries in degrees of visual angle.

Generally, pixel-wise IoU shows consistently lower agreement between the methods than radial-weighted IoU, which is expected since mismatch between the VF areas will predominantly occur near the boundaries of the VFs or in peripheral areas, which – as discussed in the previous chapter - are weighted more heavily in the pixel-wise IoU calculation than in the radial-weighted variant.

Since the duration of the PVFM measurement is almost directly proportional to the number of samples acquired per set, it is of practical interest whether smaller sample sizes than the ones captured in this experiment are already sufficient to achieve comparable agreement with kinetic perimetry. If so, the number of samples per measurement — and thus the overall measurement duration — could be substantially reduced without a meaningful loss in accuracy. Figure 5 illustrates how the number of PVFM samples used affects agreement with the kinetic perimetry, represented by radial-weighted IoU. The IoU increased most steeply between 50% and 80% of the total sample count (reaching from approximately 180 to 300 samples per measurement, or 15 to 25 samples per meridian). Beyond this range, additional samples resulted in considerably smaller average improvement in IoU, suggesting a saturation effect.

**Fig 5.**
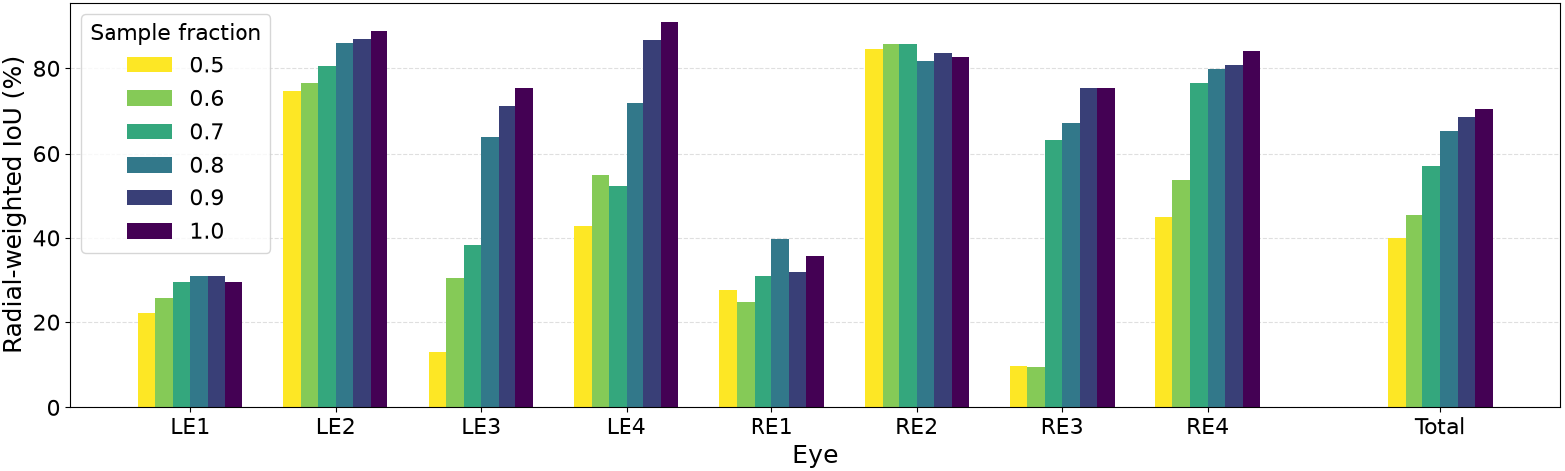
PVFM accuracy based on number of samples. Agreement of kinetic perimetry based visual fields and visual fields based on different sub-sets of the PVFM samples, shown as the radial-weighted Intersection over Union (IoU) between both methods. The sample fraction describes the amount of samples from the PVFM measurements (∼360-400 samples per set) that are used to calculate the visual field; i.e. 0.5 means that only the first 50% of samples were considered for this calculation, simulating a shorter measurement session.

### Questionnaires

The questionnaire concluding each patient session (Figure 6) was aimed at evaluating the patients’ subjective perception of the two VF measurement methods and their preferences between them based on different factors.

**Fig 6.**
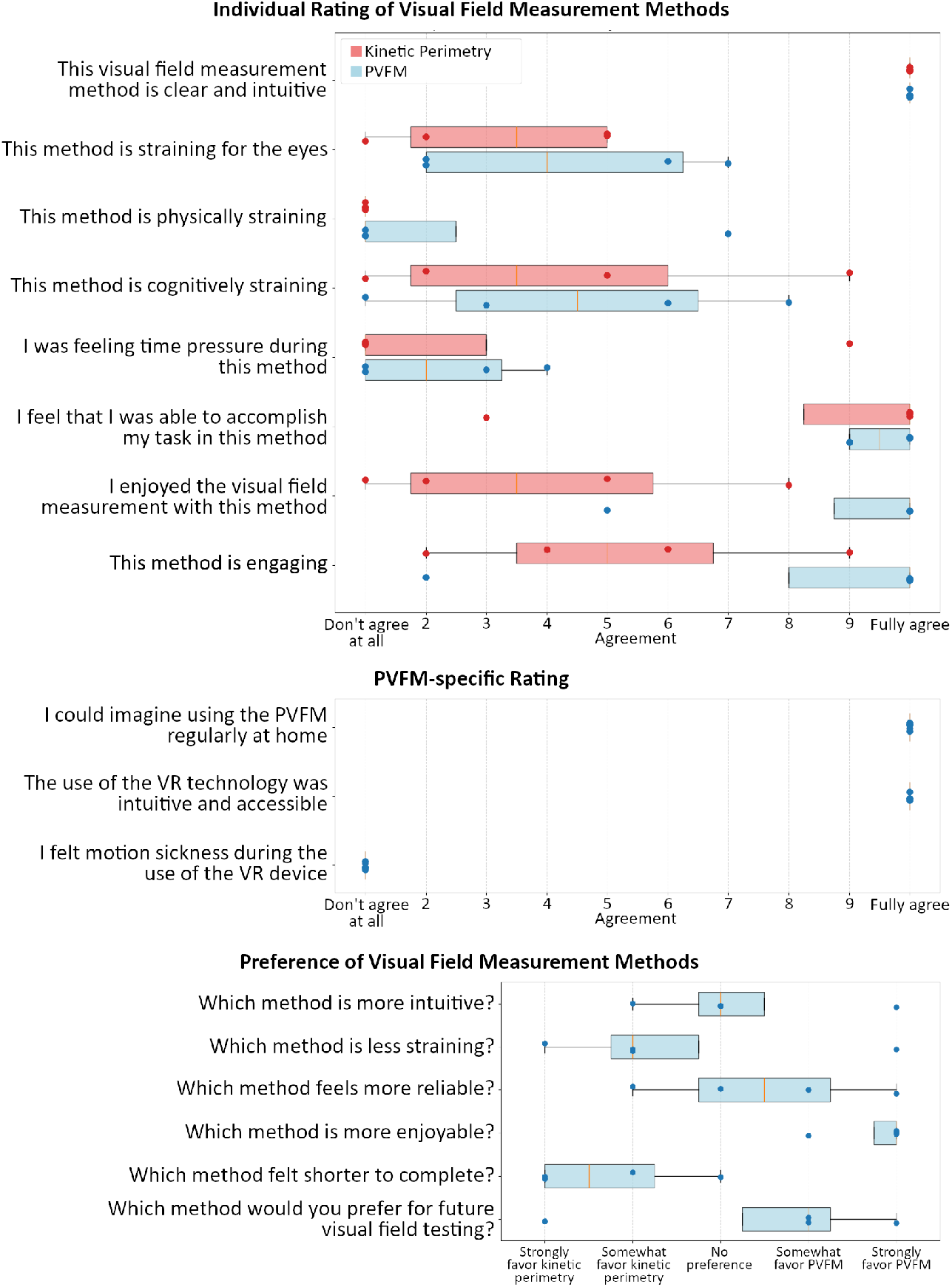
Questionnaire results. Results of the final questionnaire, showing patients’ subjective experience with kinetic perimetry and Passive Visual Field measurement (PVFM). Orange lines show the median, box boundaries show interquartile range (IQR) from 25^th^ and 75^th^ percentile, and whiskers show range of the furthest sample within 1.5 · IQR distance.

Both methods were unanimously rated as highly intuitive (median: 10/10, range: 10–10 for both). Patients reported moderate cognitive strain (kinetic perimetry: median 3.5/10, range 1–9; PVFM: median 4.5/10, range 1–8) and mild-to-moderate eye strain (kinetic perimetry: median 3.5/10, range 1–5; PVFM: median 4/10, range 2–7). The most pronounced differences between the two methods are found for enjoyment and engagement. PVFM received noticeably higher median scores for both enjoyment (10/10, range 5–10 vs. 3.5/10, range 1–8 for kinetic perimetry) and engagement (10/10, range 2–9 vs. 5/10, range 2–9 for kinetic perimetry). Additionally, the variation in patient ratings was found to be quite substantial across both conditions (mean score range of 4.4 for kinetic perimetry and 5.1 for PVFM), indicating strong individual differences in user experience.

When directly comparing specific aspects of both approaches, patients generally found kinetic perimetry slightly less straining, yet showed a clear preference for PVFM in terms of overall enjoyment. However, patients predominantly perceived the kinetic perimetry session to be shorter, which is consistent with actual timing measurements (Table 2) that confirmed that kinetic perimetry was consistently faster for all participants. Overall, three out of four patients expressed a preference (somewhat or fully) for using PVFM in future visual field assessments.

Furthermore, all four participants indicated willingness to use the PVFM tool regularly in a home-based setting, reported high ease of operating the VR system did not perceive any signs of motion sickness during the PVFM measurement process.

## Discussion

### Visual field tests

The comparison of VF boundaries (Figure 4) shows that PVFM testing yields promising results for concentric, central visual fields, which are common in late-stage retinitis pigmentosa or glaucoma. However, PVFM performs worse at detecting peripheral areas of remaining vision, showing noticeably smaller VFs than kinetic perimetry.

To put these VF results in context with existing literature, Grobbel et al. [41] report that for kinetic perimetry on the Octopus 900, test-retest variability averaged 1°-3° for the III4e stimulus setting at a speed of 5°/s — the same settings used in this study. Similarly, a study from Mönter et al. [8] found a mean test-retest variability of 1.3° using the Octopus 900 perimeter in a group of moderately advanced glaucoma patients, and Niederhauser et al. [6] report an intra-patient test-retest standard deviation of around 1° for Goldmann kinetic perimetry.

While the exact methodology of these studies differs from ours, limiting the comparability, the agreement between PVFM and kinetic perimetry for mostly concentric VFs — which ranged from 0.31° to 2.68° — falls within the range of test-retest variability reported in the literature above. This suggests that PVFM may be capable of providing VF predictions for concentric central VFs at the level of clinically conventional methods.

However, as noted earlier, one patient with a non-concentric VF showed substantial differences between methods, with mean absolute angular distances of 17.5°-22.3°, far exceeding the test-retest variability reported in the literature. Chapter Limitations and technical considerations will highlight potential reasons - and possible solutions - for this strong discrepancy in results. It should further be noted that, according to the kinetic perimetry results, only a single patient in the study population had remaining peripheral vision. Results for this VF type are therefore based on a sample size of one, and further testing is needed to better understand PVFM’s capabilities in this context and to explore possible optimizations.

It should also be noted that for any VF test, the ground truth of a patient’s visual field is unknown. New methods can only be compared against other methods such as kinetic perimetry. While these methods are well-established in the clinical context, it cannot be said with certainty how closely the measurement reflects the true visual field. Accordingly, not every deviation of the PVFM measurement from the kinetic perimetry measurement necessarily reflects a lack of accuracy in the PVFM method.

### Learnings from the acquired data

The initial configuration parameters for the PVFM measurement — including the number of samples required per meridian, the threshold-based boundary detection method, and its individual parameter values — were partly based on preliminary internal testing using simulated visual field defects of varying sizes. These simulations followed the approach of Neugebauer et al. [4], who showed that the performance of visually healthy individuals with a simulated peripheral visual field defect approximately matches that of real patients in VR-based visual tasks, including visual search. The sample data from actual patients acquired in this pilot study now allows for a more accurate and informed selection of PVFM configuration settings for future studies. The following sections discuss what was learned from this sample data regarding these configuration choices.

A major factor in the prediction of VF boundaries with the PVFM method is the boundary threshold calculation method. While post-experimental modification of evaluation methods is generally discouraged, since it risks “cherry-picking” the method best suited to the exact samples acquired and thereby falsely suggesting higher accuracy than should be expected for a different sample set, the data acquired in this pilot study nonetheless offers an opportunity to learn about potentially better-suited sample sizes and VF boundary detection methods.

A common method for threshold-based detection is the z-score method, in which a sample is considered within the threshold if its distance from the reference mean falls below a certain factor of the reference standard deviation, with typical factors being 2, 2.5, or 3 [32]. This method failed to provide good results during preliminary internal testing, and the same trend holds true for the actual experimental sample data, even when using outlier-robust methods analogous to those described in Chapter Passive Visual Field Measurement Tool. It is found that the threshold varies substantially between patients — even relative to their range of response times. This occurs because some patients show very consistent response times for samples with small angular distance (used as reference samples), while others show much higher variance in response times for those same samples. Since these values are not proportional to the overall spread in response time, they lead to over- or under-estimation of the boundary threshold. These findings underpin the choice of a variance-independent threshold calculation method.

Having thus further solidified the selection of the pre-experimentally defined threshold calculation of *t*_boundary_ = 1.5 · *robust mean* (Equation 2), a question remains how a higher or lower factor than 1.5 — resulting in a more or less sensitive VF area detection — would influence results. Again, the factor of 1.5 was chosen based on pre-experimental, simulated test samples, whereas the optimal method might differ for real patient sample data. Thus, several configuration variants of the PVFM algorithm were tested against the sample data to address this. These include varying the threshold factor to 1.33, 2.0, and 2.5. While the factor did influence the predicted VF boundaries, none of these values showed a clear improvement over the originally selected factor of 1.5. Smaller factors — i.e., lower thresholds — led to earlier detection of VF boundaries and thus smaller VFs, while larger factors — i.e., higher thresholds — led to larger VF sizes and detection of peripheral VF windows inconsistent with both kinetic perimetry and subjective interpretations of the visualized sample data, thus likely representing false-positive detections. As such, no evidence was found that adjusting the threshold factor by a fixed value would improve the quality of the PVFM method.

The tested calculation methods are not all-encompassing. Other approaches, such as different forms of cluster-based segmentation [42], also hold promise for this evaluation. However, implementing, testing, and comparing each of these methods is beyond the scope of this work, though it lends itself as a focus of future studies.

Beyond the boundary detection method, the sample data also provides insight into the sufficiency of the number of samples captured per set. As shown by the accuracy trend relative to sample count in Figure 5, a fraction of at least 0.8 — corresponding to roughly 300 samples per measurement, or about 25 samples per meridian — is recommended to achieve any meaningful result with the tested PVFM method, as further reducing the sample count may lead to a drastic reduction in accuracy. In other words, 25 samples per meridian may represent the best compromise between time and accuracy for the PVFM method, if time is a consideration. The average IoU across all measurement sets suggests a saturation curve between 50% and 100% of the samples, indicating that while increases in sample size beyond 30 samples per meridian may further improve alignment with conventional VF tests, the effect would likely be minor.

### Limitations and technical considerations

A major limitation of PVFM identified in this study is its low alignment with conventional VF testing in peripheral VF areas, based on the results of the one patient displaying such areas. Specifically, large portions of the peripheral VF detected by kinetic perimetry were not detected by PVFM. This discrepancy can be explained by two potential factors:

First, the target in kinetic perimetry is moving, whereas targets in PVFM are static. Since the human eye has specialized mechanisms for detecting motion [43], moving targets can be assumed to be easier to detect than static ones. This suggests that introducing movement into the visual representation of PVFM targets — such as jitter or small back-and-forth motion that largely preserves the relative position between targets while still triggering motion-sensitive perception — could improve detection in future implementations.

Second, the limited screen brightness of the applied VR headset - the Meta Quest Pro - at 100 cd/m^2^ did not allow for the target brightness to fully match that used in kinetic perimetry, which for the III4e condition is 318 cd/m^2^ [8]. Degenerative visual field loss, as caused by conditions such as retinitis pigmentosa or glaucoma, does not always result in complete loss of vision in affected areas. Instead, it can gradually reduce light sensitivity, such that bright stimuli remain perceivable while dimmer stimuli are no longer detected. The brightness difference between the two target stimuli may therefore have caused difference in peripheral VF area detection between the two methods. This technical limitation of VR based VF testing has been reported in other studies [44–46]. As this is an inherent hardware limitation of the VR devices used, no straightforward solution exists for PVFM within the current setup. Sauer et al. [14] tested the maximum brightness of several VR headsets and found only one, the HTC Vive, scoring notably higher than 100 cd/m^2^ at 180 cd/m^2^. However, restricting PVFM to a single VR device model would substantially reduce its flexibility and accessibility, particularly for at-home use. As such, limited display brightness remains an open challenge for the PVFM method.

### Opportunities and limitations of practical implementation

One goal of PVFM is flexibility for integration with a range of VR-based interactive tasks, ideally in a way that allows the PVFM to run as a background process without affecting the overlaying task design and without putting any restrictions on user behavior.

The main factor supporting this goal is the PVFM’s minimal input requirements. All data the method needs to predict VF boundaries — relative target position and response time — can be captured from natural scene interactions, without artificial or unintuitive constraints on the patient. Detecting and interacting with elements in a scene is an inherently natural process. PVFM does require patients to be encouraged to interact with targets as soon as they detect them, so that response time approximately reflects search duration. However, this kind of time-based challenge is already a standard feature of gamified tasks and training paradigms and is an intuitive driver - just informing a person that a task will be timed is usually enough for them to be inherently encouraged to complete the task quickly.

That said, while the user-facing demands of PVFM are minimal, successfully implementing it does impose several constraints on task design, which are listed in the following paragraphs.

The first and strongest is the requirement of randomized target positioning — or in other words, a visual-search component. With positionally fixed elements, such as a graphical user interface with static layout, response time would be prone to be influenced by users memorizing the locations of individual elements, undermining the method’s core assumption of response time reflecting visual search durations.

Second, to keep patients interacting with targets in rapid succession — which is what makes response time a valid proxy for detection speed — target interactions must remain cognitively simple, free of tasks like reading or strategic planning. This makes PVFM best suited to fast-paced, action-oriented gamified tasks, and less compatible with puzzle-based or slow, deliberative ones.

Third, to resolve visual field boundaries for each eye independently, targets must be presented to one VR display at a time, ensuring that any given target is perceived by only one eye. This is a fairly specific design requirement, since most tasks are not built with monocular presentation in mind. It does not preclude integration with most game tasks — task mechanics are typically independent of whether content is shown to one or both eyes — but it does require a non-trivial modification to the visual pipeline of most candidate tasks.

Beyond these hard requirements, several additional design choices can improve PVFM’s effectiveness without being strictly necessary. First, the number of simultaneous targets should be kept low. Although PVFM places no absolute restriction on target count, a higher target count raises the likelihood that a target will be detected and selected quickly even when it falls outside a patient’s true VF range from a prior interaction. This narrows the response-time gap between within-VF and outside-VF samples, making boundary estimation less reliable. Second, although PVFM can theoretically accommodate target movement, the distance a target travels between initial detection and interaction — i.e., over the course of the reaction time — needs to be accounted for in the boundary detection process. Third, the visual presentation of the scene, including both target appearance and background, should remain consistent across measurements. Low target–background contrast or visually cluttered backgrounds can substantially distort boundary predictions.

PVFM was designed specifically to detect VF boundaries in peripheral visual field loss, as seen in conditions such as retinitis pigmentosa or glaucoma. Its applicability to other VF defect patterns — such as hemianopia (one-sided visual field loss) or age-related macular degeneration (central visual field loss) — has not been evaluated in this work. While the underlying response-time-based threshold detection may generalize independently of VF shape, adapting it to these patterns would likely require modifications to the threshold-calculation method, and would also affect which visual tasks are appropriate for PVFM integration.

A last consideration for practical implementation of PVFM is the time factor. The findings of this pilot study suggest that the PVFM method takes a considerable amount of time, 10 or even 20 minutes per eye. As shown in Figure 5 and discussed in Chapter , a reduction in sample size is not advisable, as it would likely decrease the accuracy of the measurement. However, unlike kinetic perimetry and other “active” VF measurement methods, PVFM can potentially be embedded in tasks that patients are already motivated to perform, such as rehabilitation or preventive training exercises. In that case, the samples needed for VF prediction could be collected in the background, at no additional time cost to the user. The duration of PVFM therefore remains a design challenge for future work, but it becomes a minor concern if the method is integrated into an existing task environment.

## Conclusion

In this pilot study, we investigated the feasibility of predicting the visual field of patients with peripheral visual field defect through natural interactions in a virtual reality based visual task, without restricting the patients’ gaze behavior or forcing other non-intuitive behavior. The developed Passive Visual Field Measurement method was capable of predicting visual field boundaries of mostly concentric visual fields with an accuracy of 0.32° to 2.68° mean angular distance compared to conventional automated kinetic perimetry testing, approximately matching the test-retest variance of those clinical visual field tests. However, for peripheral visual field areas of non-concentric visual fields, the developed method shows weak alignment with the kinetic perimetry reference. Overall, while the small study population (n=4) mandates further testing to statistically validate our findings, these preliminary results suggest that Passive Visual Field Measurement is a promising, accessible, intuitive, and engaging virtual reality based tool for monitoring peripheral visual field defects. However, improving detection in peripheral visual field areas remains a key challenge for future work.

## Supporting information

Supporting File 1 - Passive Visual Field Measurement samples

Supporting File 2 - Questionnaire results

## Data Availability

All data are included in this manuscript in the form of supporting information files

## Supporting information

**S1 File. Passive Visual Field Measurement samples**. A CSV file containing all samples of the PVFM session.

**S2 File. Questionnaire Results**. A CSV file containing the questions and all participants’ answers from the final questionnaire.

## Notes

### Competing Interest Statement

The authors have declared no competing interest.

### Author Declarations

The study was approved by the ethics committee of the faculty of medicine and the university hospital of the University of Tuebingen (Nr. 701/2024BO1) in accordance with the 2013 Helsinki declaration. Patients were recruited and measurements were taken in the time from May 1 to May 31, 2026. All participants received and signed written informed consent forms.

### Funding

- This work was funded by the Federal Ministry of Education and Research (BMBF) and the Baden-Württemberg Ministry of Science as part of the Excellence Strategy of the German Federal and State Governments.

